# Influenza Virus Genomic Surveillance, Arizona, USA, 2023-2024

**DOI:** 10.1101/2024.02.26.24303283

**Authors:** Rabia Maqsood, Matthew F. Smith, LaRinda A. Holland, Regan A. Sullins, Steven. C. Holland, Michelle Tan, Gabrielle M. Hernandez Barrera, Alexis W. Thomas, Mario Islas, Joanna L. Kramer, Lora Nordstrom, Mary Mulrow, Michael White, Vel Murugan, Efrem S. Lim

**Author notes:** These senior authors contributed equally to this article (Vel Murugan, Efrem S. Lim). Address for correspondence: Vel Murugan, Arizona State University, PO Box 876101, Tempe, AZ 85287, USA; Efrem S. Lim, Arizona State University, PO, Box 876101, Tempe, AZ 85287, USA;.

## Abstract

We conducted genomic sequencing of 110 influenza A and 30 influenza B viruses from specimens collected between October 2023 and February 2024 in Arizona, USA. We identified mutations in the hemagglutinin (HA) antigenic sites. Real-time genomic surveillance is important to ensure influenza vaccine effectiveness.

## Research

Influenza viruses are common causes of acute respiratory infections that result in significant morbidity and mortality annually (1). Continued selection of amino acid mutations drives the evolution of influenza virus to undergo antigenic drift (2). As a result, virus lineages selected for influenza vaccines must be regularly updated for optimal vaccine effectiveness (3). Genomic surveillance of contemporary influenza viruses in real-time plays an important role for public health. In the prior 2022-2023 season, influenza A(H3N2) virus was the dominant virus subtype that circulated in the United States, over influenza A(H1N1)pdm09 and influenza B/Victoria (4).

To monitor influenza viruses circulating in Arizona, USA, we conducted genomic sequencing of influenza viruses in the 2023-2024 season (**Figure 1A**). As part of ongoing vaccine effectiveness studies, symptomatic individuals seeking care for an acute respiratory infection of ≤ 7 days duration at ambulatory settings in Arizona (Arizona State University’s Health Services, Phoenix Children’s Hospital and Valleywise Health) were recruited. Respiratory specimens (nasopharyngeal swabs and throat swabs combined) were tested for SARS-CoV-2, influenza A and influenza B viruses using real-time RT-PCR (RT-qPCR) multiplex assays (Applied Biosystems TaqPath COVID-19, FluA, FluB Combo Kit). Influenza-positive specimens with RT-qPCR assay cycle threshold values of less than 30 were selected for whole genome sequencing. Next-generation sequencing (Illumina, 2 × 150 paired end) was performed using cDNA amplification with termini “Uni” primers (5) and/or hybrid-capture enrichment (Illumina Respiratory Virus Oligo Panel v2). The average sequencing depth was 10.6 million paired end reads per specimen. Sequencing reads were trimmed with Trim Galore version 0.6.10 (6) and mapped against a custom database of influenza reference genome sequences. Mapped reads were used to assemble consensus sequences for each gene segment with a minimum depth threshold of 10 and quality filter of 20. We assembled the complete/near complete genome sequences of 110 influenza A virus and 30 influenza B virus (GISAID Accessions: EPI_ISL_18928538, EPI_ISL_18928541, EPI_ISL_18930210-18930347). The median age among sequenced cases was 21 (range 0.9 - 74) years; 66 (47.1%) were male and 74 (52.9%) were female; 58 (41.4%) identified as Hispanic or Latino, 76 (54.3%) as not Hispanic or Latino, and 6 (4.3%) responded with “Don’t know” or “Prefer not to answer”. Specimen collection dates ranged from October 12^th^, 2023 to February 5^th^, 2024. Three specimens with co-infections of influenza A(H1N1)pdm09 and influenza A(H3N2) were omitted from the analyses.

**Figure 1.**
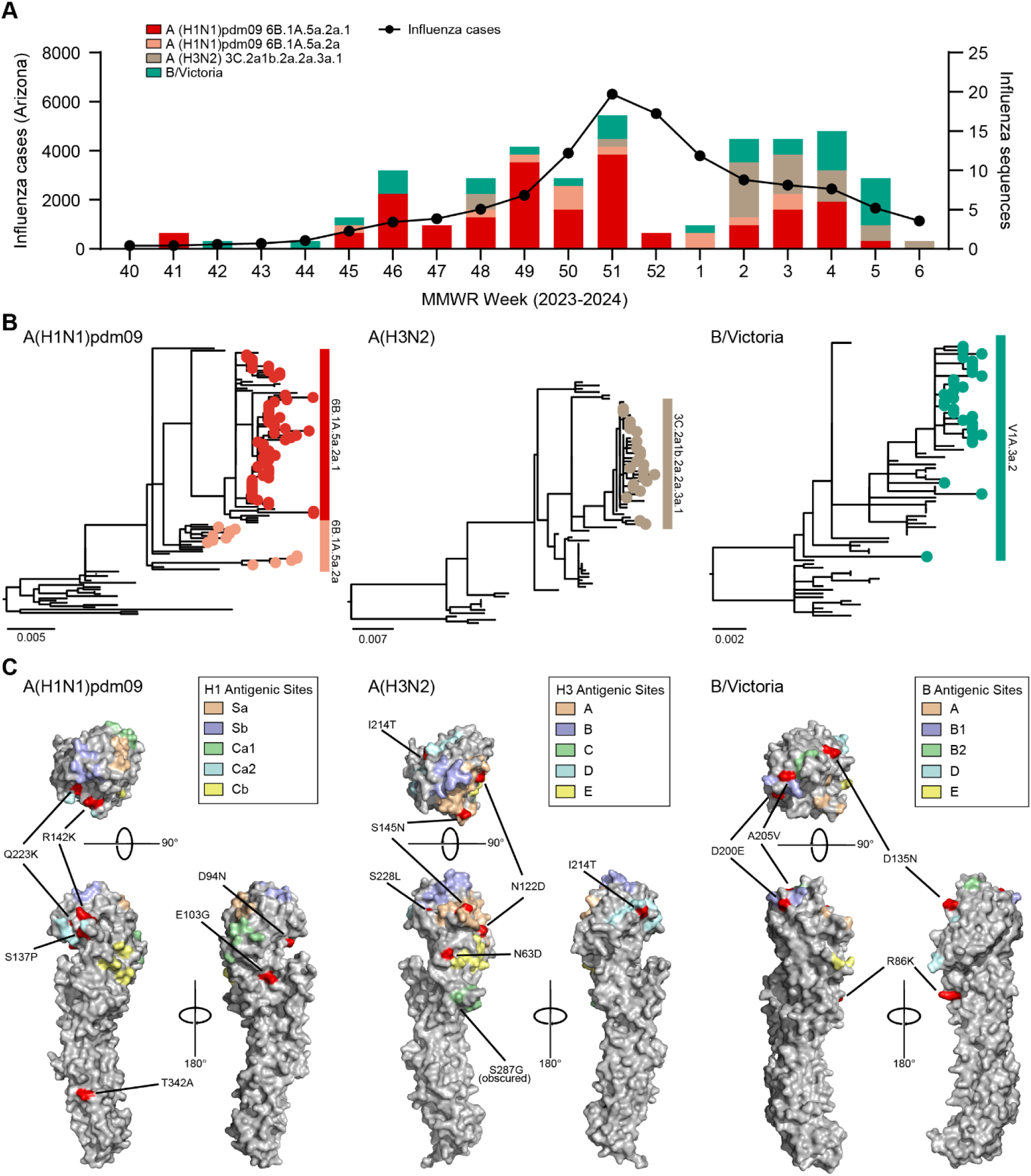
Genomic sequencing analysis of influenza viruses in Arizona, USA, 2023-2024. A) Five-week moving average of laboratory-confirmed influenza cases in Arizona reported by the Arizona Department of Health Services and influenza genome sequence counts by subtype/clade obtained for specimens used in this study. B) Phylogenies of influenza A(H1N1)pdm09, A(H3N2) and B/Victoria viruses HA gene is shown. Arizona sequences from this study are indicated. C) Structure of influenza HA proteins and locations of important residues. Mutations predicted by FluSurver as interest levels 2 and 3 (red) and antigenic sites (shaded color regions) were mapped to representative crystal structures for A(H1N1)pdm09, A(H3N2), and B/Victoria proteins (PDB: 7KNA, 4WE8, 4FQM). Models were visualized and manipulated using PyMOL.

Phylogenetic analyses of HA gene (n=128) showed that the Arizona sequences were primarily influenza A(H1N1)pdm09 clades 6B.1A.5a.2a.1 (48.4%, n=62) and 6B.1A.5a.2a (10.2%, n=13), A(H3N2) clade 3C.2a1b.2a.2a.3a.1 (18%, n=23), and B/Victoria clade V1A.3a.2 (23.4%, n=30) (**Figure 1B**). We did not detect influenza B/Yamagata lineages. Based on their vaccination status in the time since July 1^st^, 2023: 119 (85.0%) of sequenced cases reported not having received an influenza or COVID-19 vaccination, while 8 (5.7%) report having received both vaccinations, 8 (5.7%) report having received only COVID-19 vaccination, 3 (2.1%) report having received only an influenza vaccination, and 2 (1.4%) reported that they did not know whether they had received either vaccination. Of the influenza vaccinated individuals, 8 were infected with influenza A(H1N1)pdm09 clade 6B.1A.5a.2a.1, 1 influenza A(H3N2) clade 3C.2a1b.2a.2a.3a.1, and 2 influenza B/Victoria clade V1A.3a.2. The most frequent symptoms reported were fatigue (89.3%, n=125), runny nose or nasal congestion (88.6%, n=124), and fever or feverishness (84.3%, n=118). One individual had a coinfection of influenza A(H1N1)pdm09 clade 6B.1A.5a.2a.1 and SARS-CoV-2 lineage JN.1.

We identified 56 non-synonymous mutations in A(H1N1)pdm09 HA gene sequences, 33 in A(H3N2) and 18 in B/Victoria when comparing to current 2023-24 WHO cell culture and recombinant-based influenza vaccine strains. Of these mutations, 6 A(H1N1)pdm09, 6 A(H3N2) and 5 B/Victoria mutations were predicted to be of high phenotypic consequence (FluSurver tool (7) interest levels 2 and 3; **Appendix Tables 1–3**). We mapped these mutations of interest to the antigenic sites (as defined in (8)) overlaid onto influenza HA protein crystal structures (PDB: 7KNA, 4WE8, 4FQM; **Figure 1C**). The mutations were surface exposed, and many were in, or adjacent to, known antigenic sites (**Appendix Tables 1–3**). There were no mutations uniquely conserved in vaccine breakthrough cases. One non-synonymous mutation S247N in A(H1N1)pdm09 neuraminidase sequences (4.1%) was predicted to have potential antiviral impact.

Our genomic surveillance shows that the influenza viruses circulating in the 2023-24 season shifted towards the A(H1N1)pdm09 subtype in Arizona, USA, as compared to the prior season. Our findings highlight the continued adaptive evolution of influenza viruses. A limitation of this study is the sequencing selection criteria of specimens with RT-qPCR assay cycle threshold values of less than 30 may bias towards influenza infections with high viral load. Whether the changes identified in HA antigenic site regions are functional, immune escape adaptive mutations will need to be addressed in future studies. Overall, this study demonstrates the importance of genomic sequencing surveillance for clinical and public health decisions.

## Data Availability

All data produced in the present study are available upon reasonable request to the authors.

## Acknowledgements

We thank the authors from originating laboratories responsible for obtaining the specimens and the submitting laboratories where genetic sequence data were generated and shared via the GISAID initiative.

This study was approved by site reliance on Duke University (Pro00112115) and Arizona State University (STUDY00011967) Institutional Review Boards and was supported in part by Arizona State University, and the Centers for Disease Control and Prevention (CDC U01 IP001180).

## Competing interests

The authors declare no competing interests.

## Author Bio

Rabia Maqsood is a Bioinformatics Analyst at Arizona State University under the supervision of Dr. Efrem Lim. Her research interests are in the human virome and pathogen genomics.

## Contributions

Conceptualization: E.S.L.; Formal analysis: R.M., M.F.S.; Investigation: R.M., M.F.S., R.A.S., S.C.H., M.T., G.M.H.B., E.S.L.; Resources: M.I., J.L.K., L.N., M.M., M.W., V.M.; Data curation: R.M., M.F.S., L.A.H., A.W.T.; Writing-original draft: R.M., M.F.S., S.C.H., E.S.L.; Writing-review and editing: R.M., E.S.L.; Supervision: E.S.L.; Funding acquisition: V.M., E.S.L. All authors reviewed and approved the final manuscript.

## Appendix

**Appendix Table 1:** Frequency of non-synonymous amino acid substitutions in influenza A(H1N1)pdm09 virus HA gene found in Arizona genome sequences compared to the 2023-24 WHO cell culture and recombinant-based vaccine reference Influenza A/Wisconsin/67/2022(H1N1)pdm09 (OQ203982), according to the recommended numbering scheme for Influenza A HA Subtypes (9).

| Mutation | Frequency | Antigenic site |
| --- | --- | --- |
| D35E | 1.40% |  |
| D35Y | 1.30% |  |
| N38D | 2.70% |  |
| R45K | 43.20% |  |
| V47I | 2.70% |  |
| E68K | 1.30% |  |
| S69P | 2.70% |  |
| A73T | 1.40% |  |
| R74I | 1.40% |  |
| N84S | 2.70% |  |
| S85P | 1.30% |  |
| D94N | 6.70% | - |
| I96T | 1.40% |  |
| I96V | 1.30% |  |
| E103G | 1.40% | - |
| R113K | 16.00% |  |
| I116M | 1.40% |  |
| T120A | 16.20% |  |
| N130K | 1.30% |  |
| S137P | 16.00% | Ca2 |
| R142K | 12.20% | Ca2 |
| F144L | 1.40% |  |
| K146E | 1.40% |  |
| K169Q | 4.10% |  |
| G170E | 1.40% |  |
| A216T | 14.10% |  |
| Q223K | 13.80% | Ca2 |
| R223Q | 87.80% |  |
| E260D | 17.30% |  |
| A261T | 1.30% |  |
| A261V | 1.30% |  |
| A277T | 18.70% |  |
| I286M | 1.30% |  |
| V295I | 1.30% |  |
| K302E | 8.00% |  |
| S309R | 1.30% |  |
| T342A | 2.70% | - |
| D356E | 1.30% |  |
| D356N | 1.30% |  |
| I404V | 1.30% |  |
| F415L | 1.30% |  |
| V418I | 6.70% |  |
| V427I | 17.30% |  |
| T434I | 1.30% |  |
| D436N | 1.30% |  |
| R450K | 1.30% |  |
| H451N | 17.30% |  |
| K454R | 1.30% |  |
| K480R | 4.00% |  |
| D501N | 1.30% |  |
| I510V | 1.30% |  |
| L526M | 1.30% |  |
| I533V | 6.70% |  |

**Appendix Table 2:**
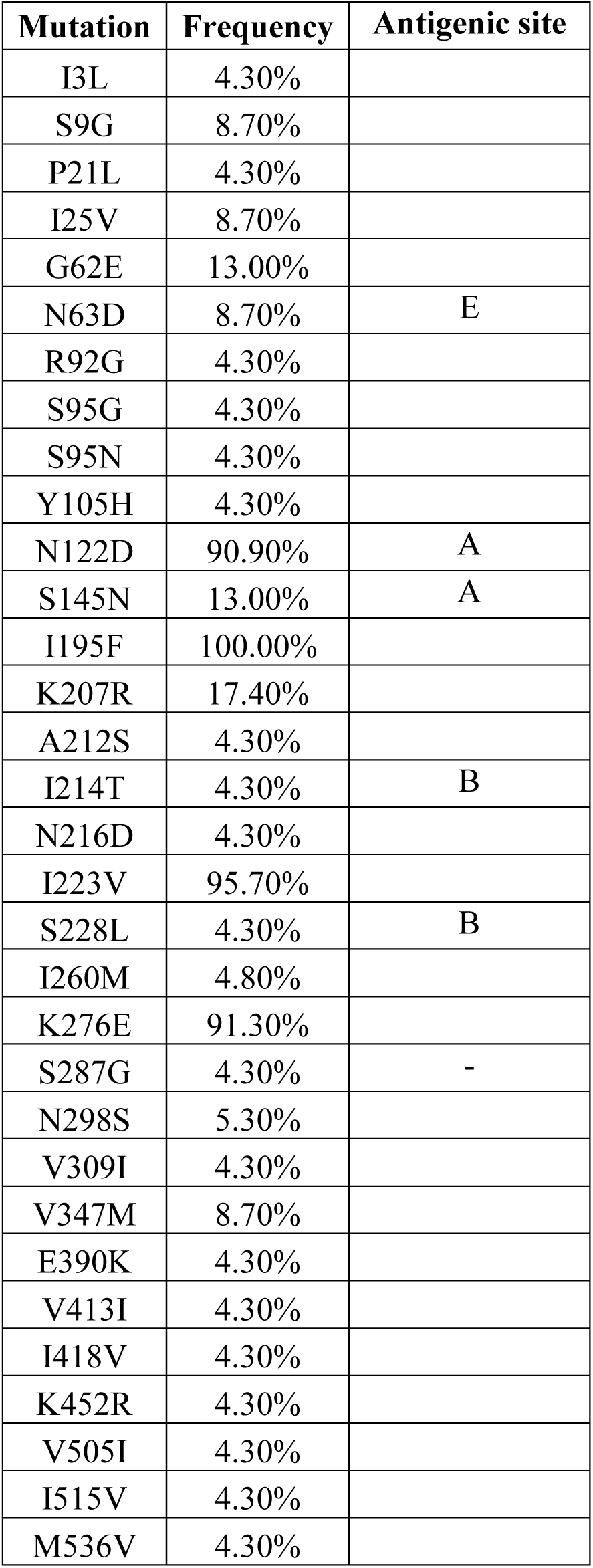
Frequency of non-synonymous amino acid substitutions in influenza A(H3N2) virus HA gene found in Arizona genome sequences compared to the 2023-24 WHO cell culture and recombinant-based vaccine reference Influenza A/Darwin/6/2021(H3N2) (OQ718999), according to the recommended numbering scheme for Influenza A HA Subtypes (9).

**Appendix Table 3:** Frequency of non-synonymous amino acid substitutions in influenza B/Victoria virus HA gene found in Arizona genome sequences compared to the 2023-24 WHO cell culture and recombinant-based WHO vaccine reference Influenza B/Austria/1359417/2021 (B/Victoria) (EPI_ISL_2378894).

| **Mutation** | **Frequency** | **Antigenic site** |
| --- | --- | --- |
| V30I | 3.30% |  |
| R86K | 3.30% | - |
| V93A | 10.00% |  |
| E134G | 3.30% |  |
| E134K | 33.30% |  |
| D135N | 6.70% | - |
| M162L | 13.30% |  |
| T185A | 3.30% |  |
| E186K | 93.30% |  |
| D200E | 100.00% | B1 |
| E201G | 3.40% |  |
| E201K | 17.20% |  |
| A205V | 10.30% | B1 |
| V255M | 3.30% |  |
| W277R | 10.30% |  |
| R501K | 3.30% |  |
| N534H | 3.30% |  |

## Notes

### Competing Interest Statement

The authors have declared no competing interest.

### Funding Statement

This study was supported in part by Arizona State University, and the Centers for Disease Control and Prevention (CDC U01 IP001180).

### Author Declarations

This study was approved by site reliance on Duke University (Pro00112115) and Arizona State University (STUDY00011967) Institutional Review Boards.

### Summary of Updates

Hemagglutinin (HA) mutation amino acid positions listed in Figure 1C and Appendix Tables were revised in accordance with recommended numbering scheme for influenza A HA subtypes; Appendix Tables were updated to clarify WHO vaccine strain references.

